# Higher polygenic risk scores for schizophrenia may be suggestive of treatment non-response in major depressive disorder

**DOI:** 10.1101/2020.01.15.20017699

**Authors:** Giuseppe Fanelli, Francesco Benedetti, Siegfried Kasper, Alexander Kautzky, Joseph Zohar, Daniel Souery, Stuart Montgomery, Diego Albani, Panagiotis Ferentinos, Dan Rujescu, Julien Mendlewicz, Alessandro Serretti, Chiara Fabbri

## Abstract

**Background:** Up to 60% of patients with major depressive disorder (MDD) do not respond to the first treatment with antidepressants. Response to antidepressants is a polygenic trait, although its underpinning genetics has not been fully clarified. This study aimed to investigate if Polygenic Risk Scores (PRSs) for major psychiatric disorders and neuroticism were associated with non-response or resistance to antidepressants in MDD.

**Methods:** PRSs for bipolar disorder, MDD, neuroticism, and schizophrenia (SCZ) were computed in 1148 MDD patients recruited by the European Group for the Study of Resistant Depression. Summary statistics from largest meta-analyses of genome-wide association studies were used as base data. Patients were classified as responders, non-responders to one treatment, non-responders to two or more treatments (treatment-resistant depression or TRD). Regression analyses were adjusted for population stratification and recruitment sites.

**Results:** PRSs did not predict either non-response or TRD after Bonferroni correction. However, SCZ-PRS was nominally associated with non-response (p=0.003). Patients in the highest SCZ-PRS quintile were more likely to be non-responders than those in the lowest quintile (OR=2.23, 95% CI=1.21-4.10, p=0.02). Patients in the lowest SCZ-PRS quintile showed higher response rates when they did not receive augmentation with second-generation antipsychotics (SGAs), while those in the highest SCZ-PRS quintile had a poor response independently from the treatment strategy (p=0.009).

**Conclusions:** A higher genetic liability to SCZ may reduce responsiveness to pharmacological treatment in MDD. From a clinical point of view, our results suggest that MDD patients with low SCZ-PRS do not benefit from augmentation with SGAs.

## 1. Introduction

Major depressive disorder (MDD) is a leading cause of disability worldwide, and it constitutes a serious economic burden for society (Greenberg *et al*., 2015, WHO, 2017). Up to 60% of adequately treated patients with MDD do not reach a complete response to the first antidepressant, and about one-third shows resistance, failing to respond to multiple treatments (De Carlo *et al*., 2016). Treatment-resistant depression (TRD) was reported to six-fold increase health care costs, especially due to a higher frequency of hospitalizations and accesses to psychiatric outpatient services (Crown *et al*., 2002). In addition to its impact on health care costs, TRD is associated with a higher risk of suicide, and a greater recurrence of depressive episodes over time (Mrazek *et al*., 2014). These burdensome consequences have led to a major effort to identify clinical and biological predictors of non-response and TRD, which might guide clinical decisions. In this regard, multivariate models were developed to predict treatment response using machine learning approaches. Promising results were obtained by using clinical-demographic predictors or their combination with genetic variants or multi-marker genetic scores (Kautzky *et al*., 2015, Kautzky *et al*., 2018, Niculescu *et al*., 2015), despite replication in independent samples remains the main issues of predictive modelling in this research field.

The lack of consistent replication of findings is related to the complex genetic architecture of treatment response, whose biological underpinnings have not been completely elucidated. Response to antidepressants has a relevant genetic basis (42% of its variance is attributed to common genetic variants) and it is highly polygenic (Tansey *et al*., 2013). This is one of the main reasons why candidate gene studies and genome-wide association studies (GWAS) provided mostly non-replicated or non-significant findings (Fabbri *et al*., 2019, Niitsu *et al*., 2013, Wigmore *et al*., 2019). The use of aggregated polygenic approaches provided more interesting results, hence the higher power of these methods (Fabbri *et al*., 2019). Polygenic Risk Scores (PRSs) take into account the effect of multiple SNPs across the genome and they capture the polygenic nature of treatment response, which is characterized by the conjunct effect of a number of loci, each with a small effect not reaching the genome-wide significance threshold in relatively small samples (Wray *et al*., 2007, 2008). PRSs have been effectively applied in other branches of medicine to identify patients having increased risk for multifactorial diseases. For example, PRSs were able to identify individuals having more than three-fold risk of developing breast cancer, inflammatory bowel diseases, type-2 diabetes, and cardiac diseases, increasing up to 20-fold the proportion of subjects at risk who can be identified compared to screening for rare monogenic variations (Khera *et al*., 2018). Disclosing clinical risk indices estimated using PRSs for Coronary Artery Disease to patients also led to better control of cholesterol levels through facilitated acceptance of statin therapies (Kullo *et al*., 2016). PRSs will likely become a valuable tool to stratify patients with different risk or prognosis. As genotyping costs decrease, it will be increasingly possible to include polygenic information into electronic health records, facilitating the integration with clinical data.

PRSs have also been used to test the genetic overlap between potentially related traits (Garcia-Gonzalez *et al*., 2017, Tansey *et al*., 2014, Ward *et al*., 2018). A shared genetic aetiology has been highlighted for bipolar disorder (BD), MDD, and schizophrenia (SCZ), with their risk variants showing a moderate genetic correlation (Schizophrenia Working Group of the Psychiatric Genomics, 2014, Stahl *et al*., 2019), and genetic variants conferring risk for those disorders might also have an effect on treatment response. Neuroticism (NEU) has shown moderate to high positive genetic correlation with resistance to antidepressants and depression, respectively [18, 19]. So far, PRSs for BD, MDD, and SCZ have not been associated with response to antidepressants in MDD (Garcia-Gonzalez *et al*., 2017, Tansey *et al*., 2014, Ward *et al*., 2018), while SCZ-PRS was associated with poorer antipsychotic and lithium response in SCZ and BD, respectively (International Consortium on Lithium *et al*., 2018, Zhang *et al*., 2019). NEU-PRSs were shown to have a negative effect on response to antidepressants (Amare *et al*., 2018, Ward *et al*., 2018), as well as to significantly predict MDD case-control status, explaining 1.05% of the phenotypic variance (Genetics of Personality *et al*., 2015).

This study aimed to investigate if PRSs for BD, MDD, NEU and SCZ are associated with non-response or resistance to antidepressants in patients with MDD, based on the hypothesis that the genetic susceptibility to these traits may represent a stratification factor in treatment response. Associations with these PRSs may help in stratifying MDD patients according to their risk of treatment-resistance and provide useful clinical information.

## 2. Methods

### 2.1. Target sample: European Group for the Study of Resistant Depression (GSRD)

#### 2.1.1. Sample participants

The sample included 1346 adults recruited by the European Group for the Study of Resistant Depression (GSRD) within a multicentre study. All the participants were diagnosed with MDD according to DSM-IV-TR criteria by the Mini International Neuropsychiatric Interview (MINI) (APA, 2000, Sheehan *et al*., 1998). Inclusion criteria included treatment with at least one antidepressants at an adequate dose for ≥4 weeks during the current MDD episode and a Montgomery–Åsberg Depression Rating Scale (MADRS) score >22 at the beginning of the current episode (Montgomery and Åsberg, 1979). Patients were excluded if they were diagnosed with any other primary psychiatric disorder or substance disorder in the preceding six months. MADRS was used to measure the current depressive symptom severity and that at the onset of the current MDD episode. Careful pharmacological anamnesis related to the present MDD episode, as well as clinical and socio-demographic data, were collected. Treatment with antidepressants was conducted naturalistically following best clinical practice principles.

All procedures involved in this study conform to the ethical standards of the Helsinki Declaration. All procedures were approved by the local ethics committees of each recruiting centre (coordinating centre approval number: B406201213479). Additional details can be found elsewhere (Dold *et al*., 2018). All patients included in this study provided written informed consent.

#### 2.1.2. Phenotypes

Patients were classified in responders, non-responders, and TRD. Response was defined as a MADRS score <22 and a decrease of at least 50% compared to the onset of the current MDD episode after at least four weeks of treatment. Non-responders were patients who did not respond to one antidepressant during the current episode. TRD was defined as a lack of response to at least two antidepressants of adequate duration (at least four weeks) and dose.

#### 2.1.3. Genotyping and quality control (QC)

All individuals were genotyped using the Illumina Infinium PsychArray 24 BeadChip (Illumina, Inc., San Diego) platform.

Pre-imputation QC was performed by removing SNPs with a genotype missing rate ≥5% and monomorphic variants. Participants were excluded if they had a genotyping rate <97%, sex discrepancies, abnormal heterozygosity, high relatedness (identity by descent (IBD) >0.1875) (Anderson *et al*., 2012), or if they were not of Caucasian ethnicity. Population outliers were defined as those outside five standard deviations for the first 20 population principal components obtained on linkage-disequilibrium (LD)-pruned genetic data (Patterson *et al*., 2006).

Genotype imputation was conducted using the Haplotype Reference Consortium (HRC) r1.1 2016 data as reference panel and Minimac3. After imputation, variants with poor imputation quality (*r*^2^ (estimated squared correlation between imputed genotypes and true genotypes) <0.30) (Li *et al*., 2010), minor allele frequency (MAF)<0.01 and genotype probability < 0.90 were pruned.

### 2.2. Polygenic risk scores (PRSs)

PRSs for BD, MDD, NEU and SCZ were calculated as the sum of risk alleles at trait-associated loci, weighted for the effect-sizes derived from the largest and most recent genome-wide meta-analyses of the Psychiatric Genomics Consortium (PGC-II studies) on BD, MDD and SCZ, and from the study by Baselmans et al. for NEU (Baselmans *et al*., 2019a, Howard *et al*., 2018, Schizophrenia Working Group of the Psychiatric Genomics, 2014, Stahl *et al*., 2019, Wray *et al*., 2018). Further information on base samples is reported in Table 1.

**Table 1.** PRSs prediction of non-response or resistance to antidepressants in patients with MDD from the GSRD sample. Results are shown for the best P_T_ (achieving the lowest p-value). Abbreviations: GSRD, European Group for the Study of Resistant Depression; MDD, major depressive disorder; PRSs, polygenic risk scores; P_T_, base genome-wide P-threshold; R^2^, the proportion of phenotypic variance explained; SE, standard error; SNPs, single-nucleotide polymorphisms. *Nominally significant results (p-value<0.05).

| Base trait | Base sample (sample size) | Phenotype outcome | Best $P_T$ | No. of SNPs at the best $P_T$ | PRS $R^2$ | Estimate (SE) | z-value | p-value | Empirical p-value ($p_E$) |
| --- | --- | --- | --- | --- | --- | --- | --- | --- | --- |
| Bipolar disorder | Stahl et al. 2019 (n=51,710) | response vs. non-response | 0.1 | 87,253 | 0.004 | 0.158 (0.103) | 1.539 | 0.124 | 0.399 |
|  |  | response vs. resistance | 0.2 | 143,443 | 0.003 | 0.134 (0.102) | 1.314 | 0.189 | 0.544 |
| Major depressive disorder | Wray et al., 2018 + Howard et al., 2018 (n=500,199) | response vs. non-response | 0.1 | 84,468 | 0.009 | 0.185 (0.086) | 2.150 | 0.032 * | 0.101 |
|  |  | response vs. resistance | 0.001 | 3,991 | 0.003 | 0.124 (0.083) | 1.485 | 0.137 | 0.378 |
| Schizophrenia | Ripke et al., 2014 (n=150,064) | response vs. non-response | 0.1 | 85,400 | 0.016 | 0.312 (0.105) | 2.963 | 0.003 * | 0.014 * |
|  |  | response vs. resistance | 0.1 | 85,427 | 0.001 | 0.093 (0.104) | 0.898 | 0.369 | 0.720 |
| Neuroticism | Baselmans et al., 2019 (n=523,783) | response vs. non-response | 0.5 | 92,631 | 0.002 | -0.095 (0.085) | -1.120 | 0.263 | 0.601 |
|  |  | response vs. resistance | 0.001 | 3,143 | 0.006 | 0.165 (0.084) | 1.970 | 0.049 * | 0.154 |

PRSs calculation in the target sample was performed using PRSice-2 software (Choi and O’Reilly, 2019). For genome-wide PRSs, clumping was carried out in PRSice-2 considering a r^2^ threshold of 0.1 and a window of 250 kb to remove SNPs in high linkage disequilibrium (LD) since they would have inflate PRSs (Wray *et al*., 2018). The base summary statistics were filtered according to an INFO score of 0.4. For each subject, we computed different genome-wide PRSs based on *a priori* set of eight P-value thresholds (P_T_) (i.e., 1e-4, 0.001, 0.05, 0.1, 0.2, 0.3, 0.4, 0.5), to identify the best threshold for predicting the outcomes of interest. Empirical p-values (p_E_) were calculated by performing 10k permutations in order to avoid overfitting and Type-I error (Choi and O’Reilly, 2019). Additional Bonferroni correction was applied to account for the four base psychiatric traits and two binary phenotypes tested (alpha=0.05/8=0.006). The most predicting genome-wide PRSs were decomposed by using a gene-set PRS approach to pinpoint the biological pathways that contributed most to the association. Gene-set PRSs were calculated by using the PRSet function in PRSice-2. A list of gene-sets, previously associated with base and target traits was derived from the most recent PGC genome-wide studies and GWAS Atlas (Schizophrenia Working Group of the Psychiatric Genomics, 2014, Watanabe *et al*., 2019, Wray *et al*., 2018) and downloaded from the Molecular Signatures Database (MSigDB) v7.0. The genes in each pathway were matched to their genome boundaries according to human assembly GRCh37-hg19. Clumping was performed for each gene-set separately using a window of 1 Mb. Since flanking SNPs not physically located within the gene-set region might also influence functions of the set, a cut-off (proxy threshold) of r2≥0.8 was used for gene-set membership. Pathway-based PRSs were calculated at P_T_=1 only, because gene-set PRSs containing a small portion of SNPs may be unrepresentative of the whole gene-set. For each gene-set PRS, a self-contained p-value and a competitive p-value were provided, which respectively tested the association with the target phenotype and the enrichment of signal of the specific gene-set. The competitive p-value was obtained by comparing the observed gene-set PRS association with the 10k permuted null p-value distribution of random gene-sets PRSs. Bonferroni correction, accounting for the multiple tested gene-sets, was applied (alpha=0.05/(23 tested gene-sets)=0.0022).

PRSs were used as predictors in regression models where two binary treatment outcomes were considered (response vs. non-response, response vs. TRD). Regression analyses were adjusted for population stratification and recruitment sites. The proportion of variance in non-response or resistance explained by the PRSs was estimated by Nagelkerke’s pseudo-R^2^ by the difference between the R^2^ of the full model, containing the PRS and the covariates, and the R^2^ of the null model, which contains only the covariates.

### 2.3. Power analyses

The statistical power of the PRSs at each P_T_ was calculated using the AVENGEME R-package (Dudbridge, 2013, Palla and Dudbridge, 2015). SNP-based heritability (h^2^_SNP_) estimated from the used summary statistics was 0.2 for BD, 0.1 for MDD, 0.12 for NEU, and 0.25 for SCZ (Baselmans *et al*., 2019b, Howard *et al*., 2018, Schizophrenia Working Group of the Psychiatric Genomics, 2014, Stahl *et al*., 2019). Genetic covariance between traits in the base and target datasets were hypothesized to be 25% or 50%, according to a previous study (Garcia-Gonzalez *et al*., 2017). Assuming a covariance of 50% between traits, all the analysed PRSs showed adequate power between 90% and 100%. The power of BD-PRSs decreased to a range of 18%-67% when covariance was set to 25%, while adequate power (>80%) was observed for the other PRSs.

## 3. Results

After QC, 1148 patients were included from GSRD. The clinical-demographic characteristics of this sample are shown in Supplementary Table S1. Overall, 7,605,870 variants passed QC procedures.

Regression models were adjusted for the first nine population principal components, along with recruitment centres, since they minimized the genomic inflation factor (Supplementary Fig. S1).

### 3.1. Associations between PRSs and response to antidepressants

PRSs for BD, MDD, NEU and SCZ were not associated either with non-response or resistance to antidepressants after correction for multiple testing. Summary of results for the best P_T_ (lowest p-values) is reported in Table 1.

PRSs for BD showed p-values>0.05 for all the considered P_T_ (Supplementary Figs. S2-S3).

PRSs for MDD showed nominal associations with non-response at four P_T_ (p=0.032 and p_E_=0.10 at the best P_T_), explaining between 0.7% and 0.9% of the total variance in the outcome (Supplementary Figs. S4-S5). All the nominally significant associations showed a consistent direction of effect, with higher PRSs associated with a higher probability of non-response.

NEU-PRSs did not predict either non-response or resistance to antidepressants. A weak nominal association with TRD was observed at P_T_=0.001 (p=0.049, p_E_=0.154) (Supplementary Figs. S6-S7). SCZ-PRS was nominally associated with non-response at six P_T_, explaining between 1.1% and 1.6% of the total variance in the outcome (**Fig. 1a**). The strongest association was found at P_T_ of 0.1 (p=0.003, p_E_=0.014). At P_T_=0.1, patients in the highest SCZ-PRS quintile were more likely to be non-responders than those in the lowest reference quintile (OR=2.23, 95% CI =1.21-4.10, p=0.02) (**Fig. 1b**). No pathway SCZ-PRS was associated with non-response, neither we showed enrichment for any gene-set. Nominal associations were shown for SCZ-PRSs of the gene-sets GO:0050890 (cognition; p=0.007), GO:0044309 (neuron spine; p=0.024), GO:0010975 (regulation of neuron projection development; 0.028), GO:0021953 (central nervous system neuron differentiation; p=0.034) and GO:0048667 (cell morphogenesis involved in neuron differentiation; p=0.037), explaining between 0.8% and 1.3% of the phenotypic variance (Supplementary Fig. S9; Supplementary Table S2).

**Figure 1a.**
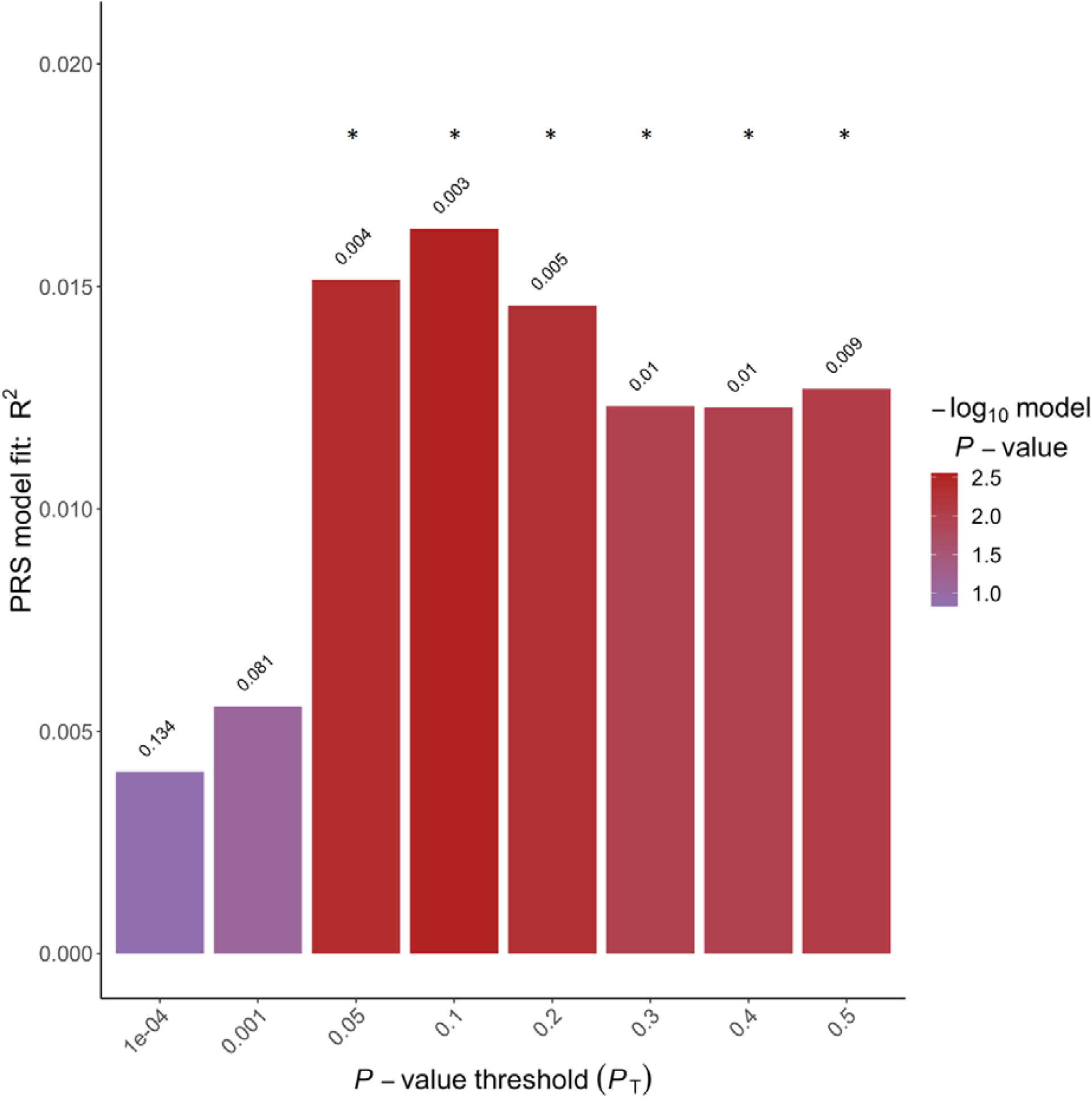
Bar plot showing the association between SCZ-PRSs and non-response to antidepressants; x-axis: GWAS p thresholds (P_T_) used for PRSs calculation; y-axis: Nagelkerke’s pseudo-R^2^. *Nominal significance level=p<0.05.

**Figure 1b.**
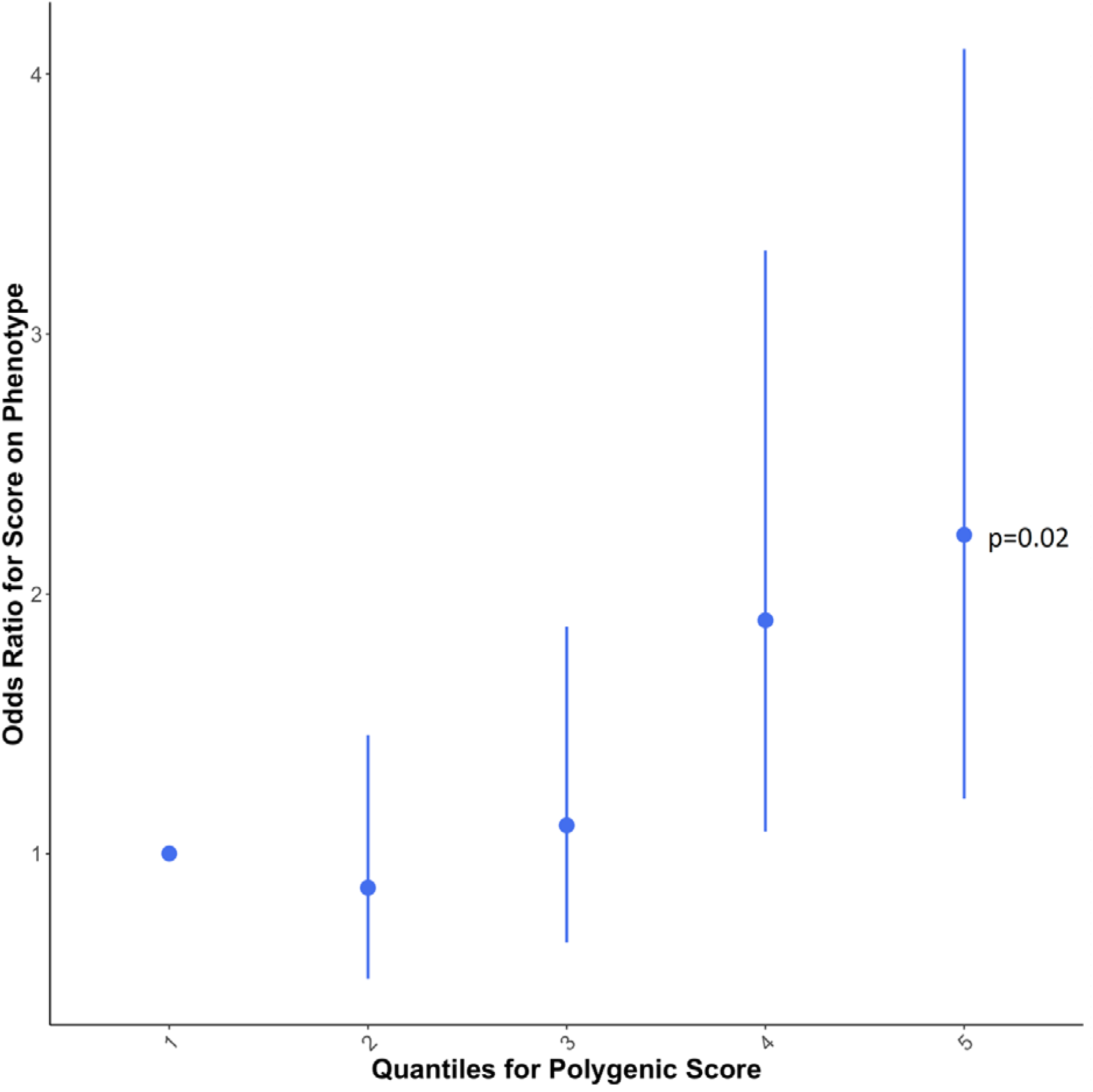
Strata plot showing the quintiles of SCZ-PRSs plotted against their effect on non-response to antidepressants. The lowest SCZ-PRS quintile was used as reference (OR=1). SCZ=schizophrenia; PRSs=polygenic risk scores; GWAS=genome-wide association study.

### 3.2. Clinical significance of the association between SCZ-PRS and non-response to antidepressants

Post-hoc univariate analyses showed that patients in the highest SCZ-PRS quintile were more frequently treated with second-generation antipsychotic (SGA) augmentation compared to patients in the lowest SCZ-PRS quintile (p=0.007). This was not due to a different distribution of psychotic symptoms between the two groups (p=0.07). The difference seems rather the consequence of a higher severity of depressive symptoms in patients in the highest SCZ-PRS quintile compared to those in the lowest quintile, in terms of current MADRS score and MADRS score at the onset of the depressive episode (p=3.5e-4 and p=0.003, respectively). Patients in the upper SCZ-PRS quintile also showed a greater number of previous depressive episodes and a more frequent family history of suicide compared to those in the lowest quintile (p=0.005 and p=0.026, respectively). Besides, they were more likely to be unemployed and to live alone (p=0.003 and p=0.028, respectively). Patients receiving augmentation with SGAs did not show higher chances of response compared to patients receiving no augmentation in either group (p=0.665 and p=0.141, respectively). Patients who did not receive augmentation with SGAs were more likely to be responders in the lowest SCZ-PRS quintile than those in the highest quintile (p=0.009) (Fig. 2), suggesting that patients in the highest quantile do not respond well to any treatment strategy and patients in the lowest quantile respond better to antidepressant monotherapy than antidepressant augmentation with SGAs. Consistently, patients treated with SGA augmentation did not show any difference in response rate depending on the SCZ-PRS quantile they belong to (p=0.972). Detailed results are reported in Table 2.

**Table 2.** Association of SCZ-PRSs with clinical and socio-demographic characteristics among responders and non-responders MDD patients from the GSRD sample. The fractions express the ratio of patients with and without the considered specific attribute. Abbreviations: SCZ, schizophrenia; PRS, Polygenic Risk Score; Q1, lowest SCZ-PRS quintile; Q5, upper SCZ-PRS quintile; SGA, second generation antipsychotics; MADRS, Montgomery–Åsberg Depression Rating Scale; MDD, major depressive disorder; BD, bipolar disorder; OCD, obsessive-compulsive disorder; PTSD, post-traumatic stress disorder. ^a^ Fisher’s exact test *p-value<0.05

|  | Lowest SCZ-PRS (Q1) | Highest SCZ-PRS (Q5) | Statistics | p-value |
| --- | --- | --- | --- | --- |
| Augmentation with SGA, no/yes | 106/28 | 85/49 | $\chi^2 = 7.289$ | 0.007 * |
| Outcome MADRS indifferently from augmentation with SGA, responders/non-responders | 62/72 | 43/91 | $\chi^2 = 5.073$ | 0.024 * |
| Outcome MADRS in patients who received augmentation with SGA, responders/non-responders | 9/19 | 18/33 | $\chi^2 = 0.001$ | 0.972 |
| Outcome MADRS in patients who did <u>not</u> receive augmentation with SGA, responders/non-responders | 53/53 | 25/58 | $\chi^2 = 6.792$ | 0.009 * |
| Psychotic features, no/yes | 93/1 | 105/7 | - | 0.073 <sup>a</sup> |
| Living with a partner, no/yes | 55/79 | 74/60 | $\chi^2 = 4.842$ | 0.028 * |
| Employment status, no/yes | 25/108 | 48/85 | $\chi^2 = 9.138$ | 0.003 * |
| Family history for MDD, no/yes | 73/50 | 75/51 | $\chi^2 = 0.01$ | 0.919 |
| Family history for BD, no/yes | 116/6 | 116/7 | - | 1.000 <sup>a</sup> |
| Any family history, no/yes | 96/31 | 96/24 | $\chi^2 = 0.462$ | 0.497 |
| Family history for SCZ-spectrum disorders, no/yes | 123/4 | 113/7 | - | 0.365 <sup>a</sup> |
| Family history for suicide, no/yes | 110/19 | 122/8 | - | 0.026 <sup>a</sup> * |
| Family history for suicide attempts, no/yes | 113/16 | 119/6 | - | 0.043 <sup>a</sup> * |
| Number of depressive episodes, mean (SD) | 3.11 (2.16) | 4.12 (3.10) | t = -2.819 | 0.005 <sup>a</sup> * |
| Age at onset (years), mean (SD) | 35.45 (15.25) | 36.22 (14.20) | t = -0.411 | 0.682 <sup>a</sup> |
| Length of hospitalisation (days), mean (SD) | 5.89 (19.06) | 5.38 (14.44) | t = 0.237 | 0.813 <sup>a</sup> |
| Melancholia, no/yes | 32/100 | 46/86 | $\chi^2 = 3.075$ | 0.079 |
| Suicidality, no/yes | 81/53 | 66/68 | $\chi^2 = 2.953$ | 0.086 |
| Agoraphobia, no/yes | 121/11 | 123/7 | - | 0.465 <sup>a</sup> |
| Social phobia, no/yes | 129/5 | 130/9 | - | 0.413 <sup>a</sup> |
| OCD, no/yes | 134/0 | 127/4 | - | 0.058 <sup>a</sup> |
| PTSD, no/yes | 133/1 | 133/1 | - | 1.000 <sup>a</sup> |
| GAD, no/yes | 122/12 | 113/21 | $\chi^2 = 2.212$ | 0.137 |
| Duration of current episode (days), mean (SD) | 166.97 (182.33) | 185.95 (175.00) | t = -0.811 | 0.418 |
| Baseline MADRS score, mean (SD) | 32.22 (7.55) | 35.08 (7.87) | t = 3.033 | 0.003 * |
| Current MADRS score, mean (SD) | 17.91 (11.18) | 22.71 (10.48) | t = -3.625 | 3.5e-4 * |

**Figure 2.**
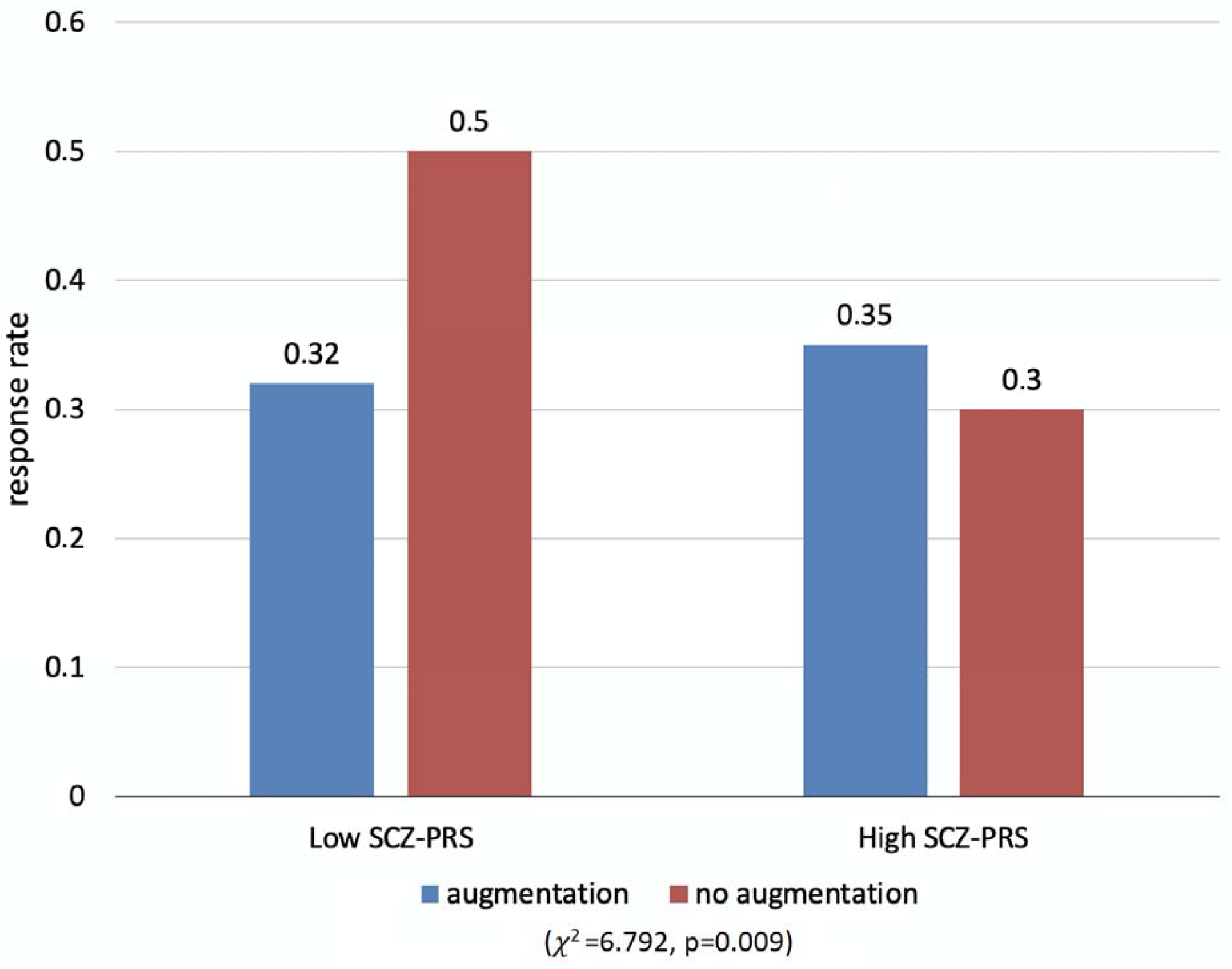
Bar plot showing the response rate of patients with MDD who receive (in blue) or not receive (in red) augmentation with antipsychotics in low SCZ-PRS and high SCZ-PRS quintile (χ2=7.89, p=0.008). MDD, major depressive disorder; antipsychotics, antipsychotics, SCZ, schizophrenia; PRS, Polygenic Risk Score.

## 4. Discussion

The present study investigated if PRSs for BD, MDD, NEU or SCZ were associated either with non-response or resistance to antidepressants in a sample of 1148 patients with MDD. Although PRSs did not show any significant effect after multiple-testing correction, non-responders to antidepressants showed trends of higher SCZ-PRSs and MDD-PRSs (p=0.003 and p=0.032, respectively). MDD patients showing higher genetic risk for SCZ were more likely to receive augmentation with antipsychotics. This clinical choice seemed driven by higher depression severity (higher MADRS score, higher number of previous episodes, higher functional impairment), rather than higher frequency of psychotic features in this group of patients. However, SGA augmentation did not improve response rates in these patients, who did not respond well to any treatment strategy. On the other hand, patients in the lowest SCZ-PRS quantile showed better response rates when they received antidepressant therapy compared to SGA augmentation, suggesting that they do not benefit or poorly tolerate this treatment. We might speculate that a higher liability to SCZ may lead to a clinically and biologically different subtype of depression, therefore less responsive to pharmacological treatments for depression. In line with this hypothesis, depressive and negative symptoms are poorly responsive to the available pharmacological treatments in SCZ (Buoli *et al*., 2016) and MDD with high SCZ-PRS was associated with less neuroticism and psychological distress and greater cognitive deficits (Whalley *et al*., 2016). Shared genetic loci between MDD and SCZ may be a starting point to identify therapeutic targets for new treatments effective in MDD with high SCZ-PRS. These loci include the major histocompatibility complex (MHC) region [27], which is involved in synaptic functionality and plasticity by modulating the activity of glutamatergic receptors and microglia-mediated synaptic pruning [36]. Glutamatergic and immune dysfunctions were linked with non-response to antidepressants, and drugs such as the Tumor Necrosis Factor (TNF)-alpha antagonist infliximab and the N-methyl-D-aspartate (NMDA)-receptor antagonist ketamine showed efficacy in MDD cases non-responsive to traditional treatments (Fond *et al*., 2014, Kim and Na, 2016). Thus, we speculate that MDD patients with higher PRSs for schizophrenia might respond better to drugs that modulate the glutamatergic and/or immune functions, but this hypothesis needs to be tested. Nevertheless, our gene-set approach was not able to demonstrate associations between any tested pathway-specific SCZ-PRSs and non-response to antidepressants and neither enrichment for the corresponding pathways. Cumulative genome-wide SCZ-PRSs, although less informative of the underpinning biology, were able to predict non-response better than the tested pathway-specific SCZ-PRSs.

To the best of our knowledge, this was the first study to indicate a suggestive overlap between SCZ genetic risk and non-response to antidepressants. Previously, in a combined target sample of ∼3700 depressed patients, no association between SCZ-PRSs and symptom improvement to antidepressants was observed, although SCZ-PRS showed a trend of association (p=0.07) and better results compared to MDD-PRS (Garcia-Gonzalez *et al*., 2017). Differently from this previous study, we investigated treatment non-response and resistance rather than considering a quantitative measure of response which is more difficult to interpret in terms of clinical significance. Our findings do not support an effect of MDD-PRS on antidepressants response, although a weak trend of association with non-response was found. Although it has not yet been replicated in adults, MDD-PRSs predicted depressive symptom severity in youths, which is a major risk factor for non-response to antidepressants (Halldorsdottir *et al*., 2019, Kautzky *et al*., 2019). Depressed patients assigned to electroconvulsive therapy, who often show a more severe and resistant phenotype, showed higher MDD-PRSs as well (Foo *et al*., 2019). Thus, a higher MDD-PRS may result in treatment non-response by the modulation of symptom severity. It is worth noting that PRSs for MDD were computed based on a GWAS meta-analysis of self-reported depressive symptoms and help-seeking behaviour, as well as clinically diagnosed MDD (Howard *et al*., 2018, Wray *et al*., 2018). It was shown that GWAS signals for minimal phenotyping-based depression might have lower specificity for MDD than those based on DSM criteria (Cai *et al*., 2018).

BD-PRSs did not predict response to antidepressants, in line with previous evidence (Tansey *et al*., 2014), while NEU-PRS showed a nominal effect on the risk of TRD, consistently with a previous GWAS (Ward *et al*., 2018), but no effect on the risk of non-response, suggesting that TRD may have a specific genetic profile. As opposed to non-response, we hypothesised that TRD might be modulated by specific genetic factors that do not significantly overlap with major psychiatric disorders, but which could be more related to personality traits.

These findings should be interpreted in light of the limitations of this study. The power of the study was estimated to be adequate for the greatest part of the performed tests, except for BD-PRS, but we did not evaluate the performance of the calculated PRSs in an independent sample or by performing cross-validation because of the size of target sample (Choi *et al*., 2018). Nevertheless, we used a permutation strategy to avoid overfitting and derive empirical p-values. The use of gene-sets is limited by current knowledge about the functioning and genes involved in the corresponding pathways. It is worth noting that PRSs consider only common genetic variants in predicting shared genetic risk between different complex traits, without considering the effect of rare variants. Finally, PRSs assume an additive effect and do not take into account any potential interactive or epistatic effect between variants.

In conclusion, this study did not identify any significant association between PRSs for three major psychiatric disorders, neuroticism and response to antidepressants, however it found a nominal association between higher SCZ-PRSs and non-response to antidepressants in MDD, an effect that should be confirmed in independent samples. If replicated, this result suggests that MDD with a higher genetic load for SCZ may represent a different biological subtype, which has lower response rates to both antidepressants and antidepressant augmentation with antipsychotics. In line with this hypothesis, patients with high SCZ-PRS showed higher symptom severity, higher disease recurrence, as well as higher evidence of functional impairment in relevant areas of life (work and intimate relationships). Patients with a low SCZ-PRS showed better response rates when treated with antidepressants only and not with SGA augmentation, suggesting that this group does not benefit from SGA augmentation but on the contrary it could cause detrimental side effects and no symptom improvement.

## Data Availability

Base summary statistics can be downloaded from https://www.med.unc.edu/pgc/results-and-downloads and https://surfdrive.surf.nl/files/index.php/s/Ow1qCDpFT421ZOO.

## Acknowledgements

We are grateful to all the participants who took part in the GSRD study and the PGC for making publicly available the GWAS summary statistics that we used as base data for our PRS analyses.

